# Association between resting-state functional brain connectivity and gene expression is altered in autism spectrum disorder

**DOI:** 10.1101/2021.01.07.21249281

**Authors:** Stefano Berto, Alex Treacher, Emre Caglayan, Danni Luo, Jillian R. Haney, Michael J. Gandal, Daniel H. Geschwind, Albert Montillo, Genevieve Konopka

## Abstract

Gene expression covaries with brain activity as measured by resting state functional MRI. However, it is unclear how genomic differences driven by disease state can affect this relationship. Here, we integrate brain gene expression datasets of neurotypical controls and autistic (ASD) patients with regionally matched brain activity measurements from fMRI datasets. We identify genes linked with brain activity whose association is disrupted in ASD patients. We identified a subset of genes that showed a differential developmental trajectory in ASD patients compared with controls. These genes are enriched in voltage-gated ion channels and inhibitory neurons, pointing to excitation-inhibition imbalance in ASD. We further assessed differences at the regional level showing that the primary visual cortex is the most affected region in ASD patients. Our results link disrupted brain expression patterns of individuals with ASD to brain activity and show developmental, cell type, and regional enrichment of activity linked genes.

## Main

### Integration of resting-state functional MRI and gene expression measures in ASD and controls

Recent advances in imaging genomics highlighted genes that covary with resting-state functional MRI (rs-fMRI) measurements across cortical regions, suggesting that gene expression might support functional signals in human brain^1-4^. However, these studies focused on neurotypical subjects, offering a window to study the effect of gene expression – functional measurement in neuropsychiatric disorders. Interestingly individuals with ASD have alterations in brain activity^5-7^ as well as gene expression patterns^8-11^. Therefore, coupling these two measurements has the potential to identify genes whose expression might promote functional networks observed in rs-fMRI and understand whether this relationship is altered in a neuropsychiatric disorder such as ASD.

Thus, to identify differentially correlated genes, we determined the spatial similarity between rs-fMRI and gene expression changes in the human brain of subjects with ASD compared to controls across 11 matched cortical regions. We used rs-fMRI data from an imaging database containing individuals with ASD and matched controls (ABIDE I^12^ and ABIDE II^13^) and cortical RNA-sequencing (RNA-seq) datasets from persons with ASD and matched controls across development into adulthood^14^ (Fig. 1). We computed two extensively validated measures of brain activation to characterize brain function from rs-fMRI. The first brain measure, fractional Amplitude of Low-Frequency Fluctuations (fALFF)^15^, quantifies the slow oscillations in brain activity that form a fundamental feature of the resting brain. The second brain measure, regional homogeneity (ReHo)^16^, a measure of local connectivity, aims to detect complementary brain activity manifested by clusters of voxels rather than single voxels as in fALFF. Voxel-wise maps of fALFF and ReHo were calculated for a total of 1983 subjects from the ABIDE I and ABIDE II datasets (ASD = 916, CTL = 1067; Extended Data Fig. 1 and Supplementary Table 1). We analyzed a total of 11 regions of interest (ROIs) matching the transcriptomic data.

**Figure 1.**
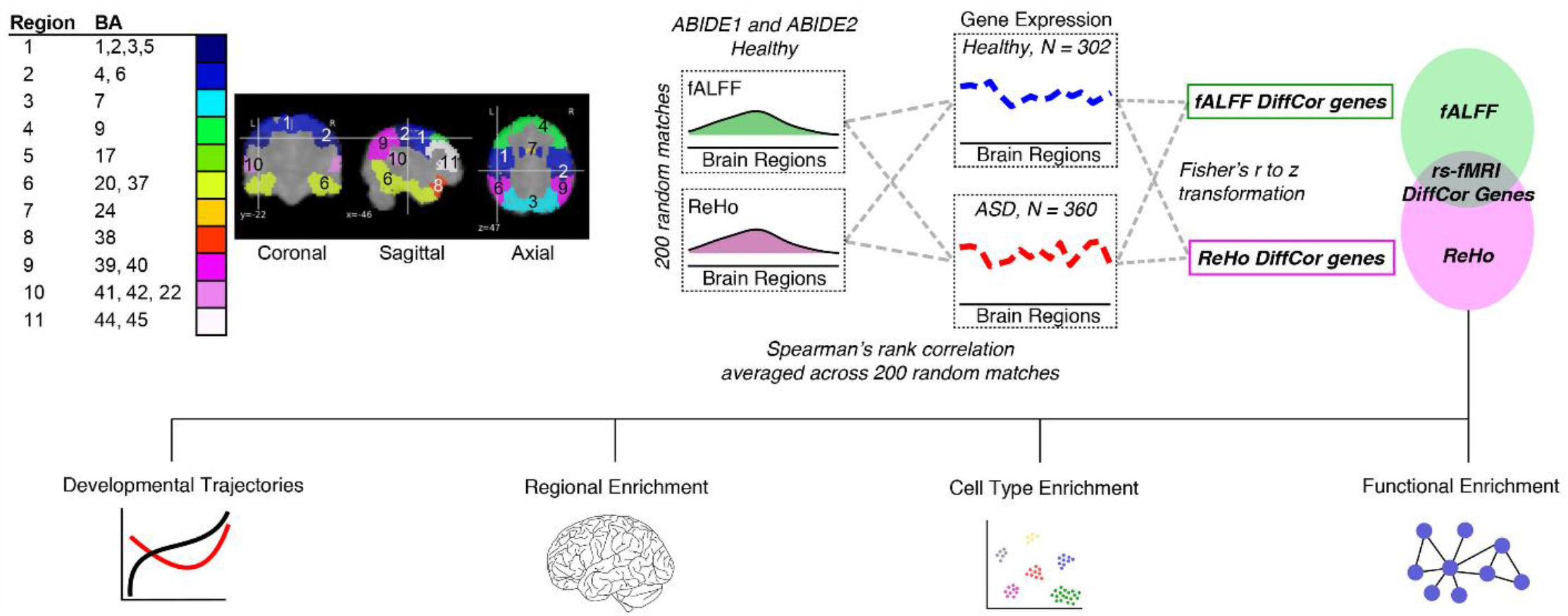
Flowchart of the analytical framework and pipeline. Gene expression values were obtained for 11 cortical regions from individuals diagnosed with autism spectrum disorder (ASD) and demographically matched neurotypical individuals (CTL: control). For fMRI, the ABIDE I and II datasets were used to select ASD and CTL individuals that demographically matched with the gene expression cohort. Regional Homogeneity (ReHo) and fractional amplitude of low frequency oscillations (fALFF) were calculated from these individuals for the 11 cortical regions. 200 subsampled matches were selected from CTL individuals. Spearman’s rank correlation was used to infer a correlation between gene expression and fMRI separately for both fMRI measurements. For differential correlation (DC genes) between ASD and CTL, the mean correlation values of matches were transformed to a z score (Fisher’s r to z transformation). A gene was called differentially correlated if the differential correlation p-value was less than 0.05 (DifCorP < 0.05) and if the gene survived FDR in CTL. This analysis was done separately for ReHo and fALFF measurements. The final list consisted of genes that are differentially correlated using both fMRI measurements.

We first assessed differences between cases and controls for both fALFF and ReHo. We identified 5 ROIs with a significant difference for fALFF and 4 ROIs for ReHo (t-test, p < 0.05; Extended Data Fig. 2a). 3 ROIs were commonly different using either measurement (BA20/37, BA3/2/5, BA7). However, we observed only a small effect size for all the ROIs analyzed (Cohen’s *d*; *d* < 0.3; Extended Data Fig. 2b) in agreement with other reports^17,18^. Since the differences between cases and controls using rs-fMRI were minimal with a small to null contribution to the analysis, we assessed the rs-fMRI – gene expression relationship using the control subject ReHo and fALFF values. We first assessed the complementarity of these two rs-fMRI measurements in controls. There was a significant correlation between fALFF and ReHo values in each singular ROI analyzed and in each cortical region (Spearman’s rho = 0.59, p < 2.2×10^−16^; Extended Data Fig. 3a,b). These data further confirmed the complementarity of these two distinct measurements of rs-fMRI values. We next took advantage of RNA-seq data^14^ from 11 cortical areas for a total of 360 tissue samples from cases (ASD) and 302 control samples (CTL) (Extended Data Fig. 4a). The variance explained by technical and biological covariates was accounted for and removed before further analyses (see Methods and Extended Data Fig. 4b).

### Identification of genes differentially correlated with rs-fMRI between ASD and controls

We sought to identify genes with correlated expression to imaging measurements across regional rs-fMRI values. To do this, we used Spearman’s rank correlation between mean regional values of fALFF or ReHo and regional gene expression. To take advantage of the entire ABIDE dataset, we randomly subsetted the ABIDE dataset 200 times and correlated each subset with the genomic data (see Methods). We defined genes correlated with ReHo and/or fALFF in both controls and ASD patients (Extended Data Fig. 5a and Supplementary Table 2). Using a Fisher r-to-z transformation, we assessed the significance of the difference between ASD and CTL correlations to define genes differentially correlated using fALFF or ReHo values (1021 and 636 respectively; Fig. 2a). We found a significant overlap between fALFF and ReHo differentially correlated genes (henceforth “DC genes”: 355; hypergeometric test, p-value = 2.75×10^−135^; Fig. 2b). DC genes showed a high proportion of positively correlated genes (70%; Extended Data Fig. 5b,c). We found that the distribution of *rho* squared confirmed a relative decreased contribution to gene expression variation by ASD individuals (Fig. 2c). We next compared the genes we identified with genes linked with rs-fMRI values from independent studies^3,4^. Because these earlier studies analyzed only healthy individuals, we first compared the genes correlated only in CTL with the ones previously reported. We found that previously fMRI-correlated genes were significantly enriched in CTL genes for both fALFF and ReHo measurements (fALFF – Wang et al.: odds ratio (OR) = 6.8, FDR = 3.3×10^−08^, fALFF – Richiardi et al.: OR = 3.4, FDR = 1.5×10^−11^; ReHo – Wang et al.: OR = 30.1, FDR = 2.2×10^−16^, fALFF – Richiardi et al.: OR = 2.8, p-value = 5.3×10^−08^; Fig. 2d). In total, 6 genes overlapped in this study and both of the previous studies, and 3 out of the 6 genes were also among the DC genes (*PVALB, SCN1B*, and *SYT2*; Fig 2e,f), revealing a certain degree of reproducibility despite variation in cortical regions and type of fMRI measurements. Among the three candidates, we found *SCN1B* - beta-1 subunit of voltage-gated sodium channel - a highly expressed gene in fast-spiking parvalbumin cortical interneurons, which play a key role in neuronal networks, and whose oscillations are linked with ASD^19-22^. Overall, these results identify many new brain activity related genes and imply that some of these high confidence genes are involved in ASD (Fig. 2e).

**Figure 2.**
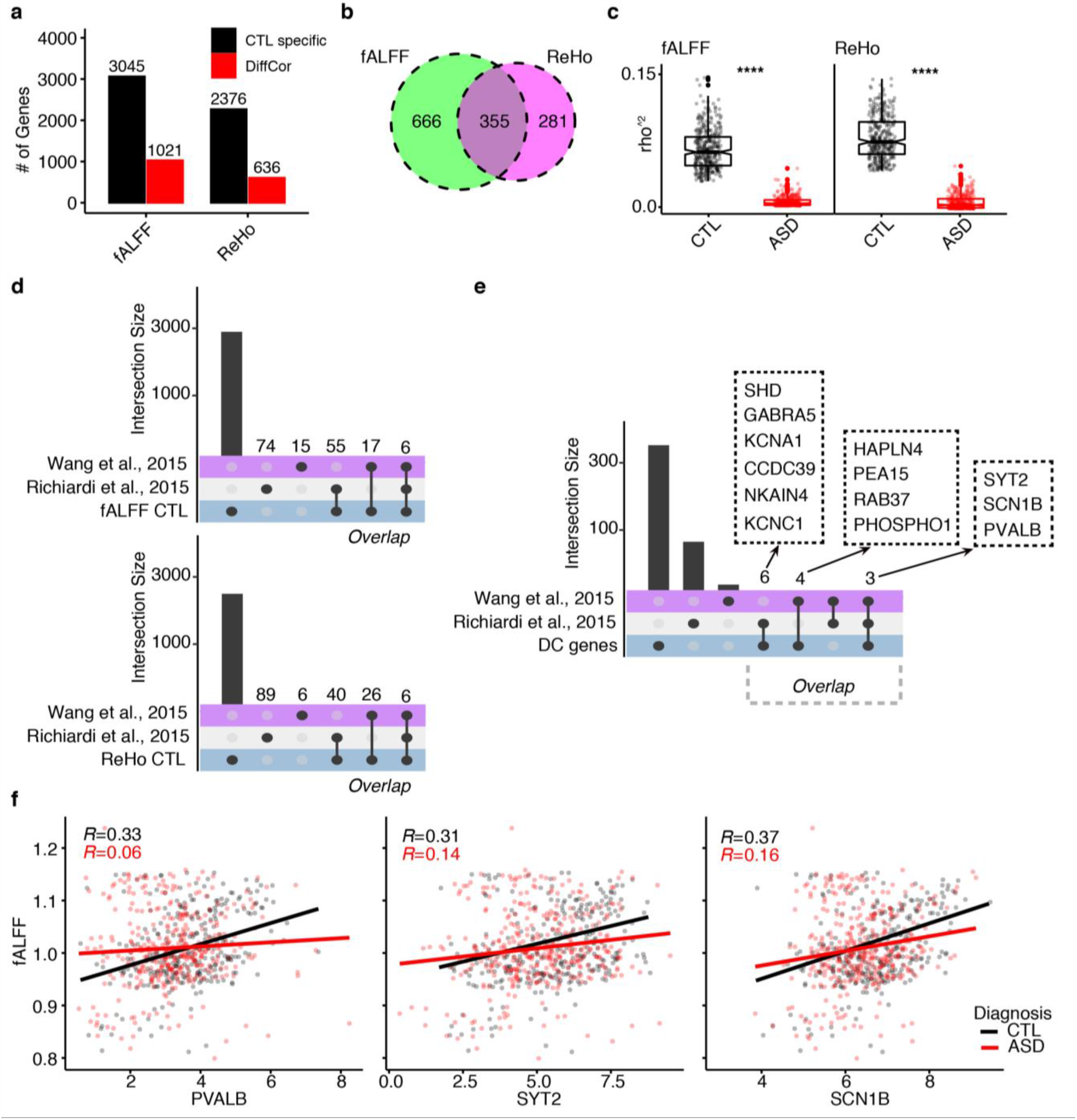
Overview of differentially correlated genes. **a**, Barplot of significant results (FDR < 0.05) in CTL (black) and the number of differentially correlated genes per each rs-fMRI measurements (red). **b**, Overlap of differentially correlated genes identified using ReHo or fALFF. **c**, Boxplot showing the differences in variance explained (Spearman’s *rho* squared) between CTL and ASD for fALFF and ReHo. Stars represent p-values from One-sided Wilcoxon’s rank-sum test (p < 0.001). Whiskers on box plots represent maximum and minimum values. Boxes extend from the 25th to the 75th percentiles, the center lines represent the median. N = 355. **d**, Upset plot showing the intersection between the genes significantly correlated only in CTL for both fALFF and ReHo and two previous rs-fMRI – gene expression studies using only data from neurotypical individuals. **e**, Upset plot showing the intersection between DC genes and two previous rs-fMRI – gene expression studies using only data from neurotypical individuals. **f**, Scatterplots showing the relationship (Spearman’s rank correlation) between rs-fMRI (Y-axis) and gene expression for three candidate genes (X-axis) in CTL and ASD.

### Differentially correlated genes have specific developmental trajectories

Although we identified DC genes across all samples with a median age of 22 years old, we asked how DC genes compare between CTL and ASD across development given that autism is a neurodevelopmental disorder. We leveraged the transcriptomic data to detect whether DC genes follow a specific developmental trajectory in ASD compared with CTL subjects (see Methods). We identified three main clusters of DC genes: one highly expressed in adults (Adult), one highly expressed in early development (EarlyDev), and one with relatively stable trajectory throughout development (Stable) (Fig. 3a). Interestingly, genes in the Adult cluster are upregulated until adulthood in neurotypical individuals but this upregulation is delayed in ASD individuals. In contrast, the genes in the Stable and EarlyDev clusters follow a similar trajectory in both groups (Fig. 4a). Because each region differs by sample size, we used a subsample approach and recalculated the developmental trajectories. We found that differences in sample size between regions did not affect the overall result (Extended Data Fig. 6a). Next, we sought to understand the functional properties of the genes associated with these developmental trajectories. The Adult cluster exhibited significant enrichment for pathways implicated in ion-channel activity (e.g. *SCN1A, SCN1B, KCNK13, KCNAB3, CACNA1S*, and *KCNA1*), while the EarlyDev and Stable clusters were enriched for neurogenesis and transcriptional activity, respectively (Fig. 3b). We also asked whether the clusters differ in their overlap with top correlated genes (TopRsq, DC genes with fALFF rho^2^ > 0.1 or ReHO rho^2^ > 0.1). Interestingly, 65% of all TopRsq genes were in the Adult cluster (Extended Data Fig. 6b), indicating that most of the top correlated genes have delayed upregulation until adulthood in persons with ASD.

**Figure 3.**
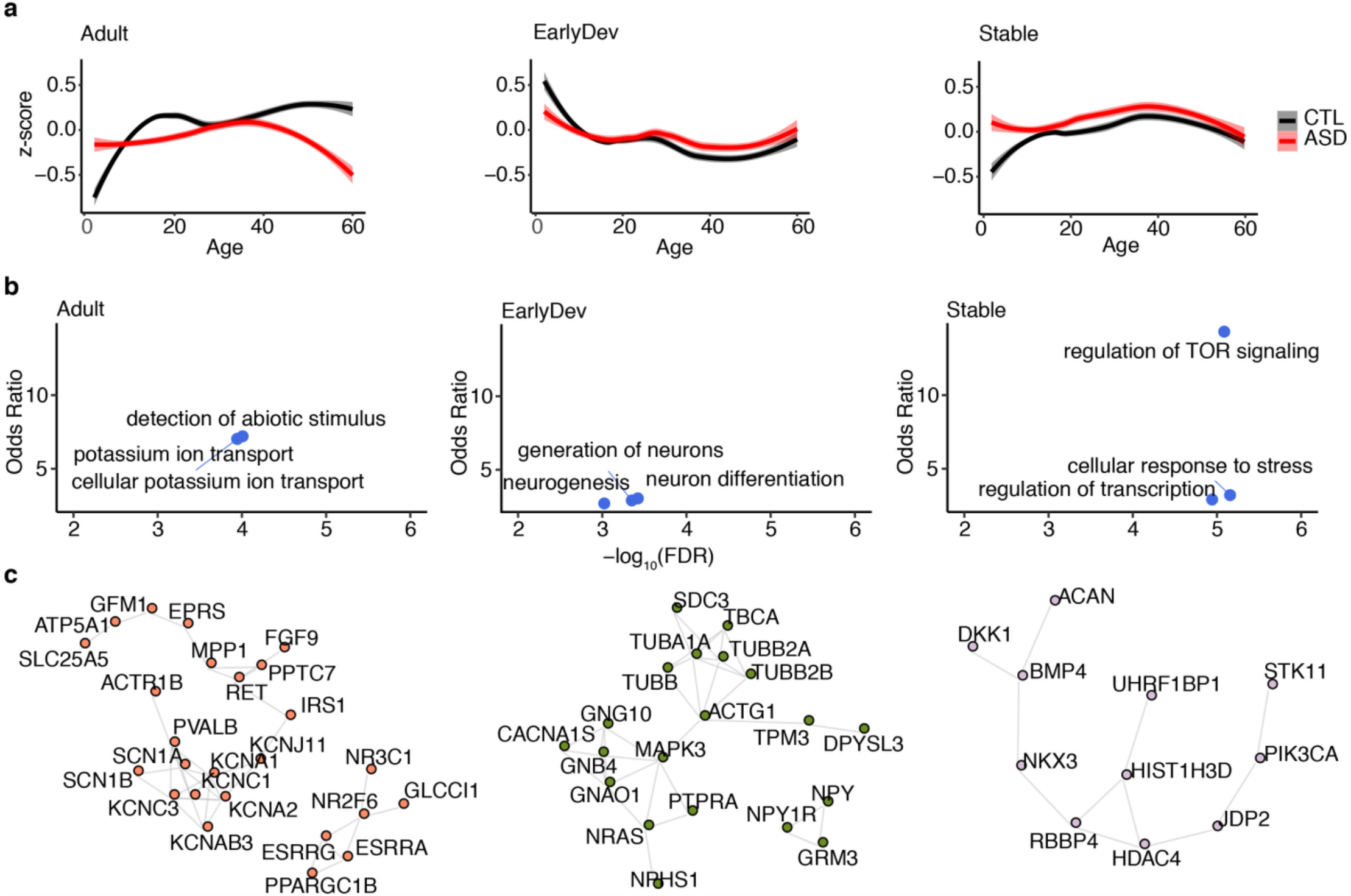
Differentially correlated genes in ASD are important for brain development. **a**, The 355 genes were clustered in three developmental time groups: adult (Adult), early development (EarlyDev), stable (Stable). X-axis represents developmental time. Y-axis represents the expression based on human brain developmental time. Loess regression with confidence intervals depicts the overall distribution. Smooth curves are shown with 95% confidence bands. **b**, Bubblechart representing the functional enrichment for modules associated with developmental time. Y-axis corresponds to the odds ratio, X-axis corresponds to the -log_10_(FDR). **c**, Protein-protein interaction based on highly scored evidence (> 300) using STRING.

**Figure 4.**
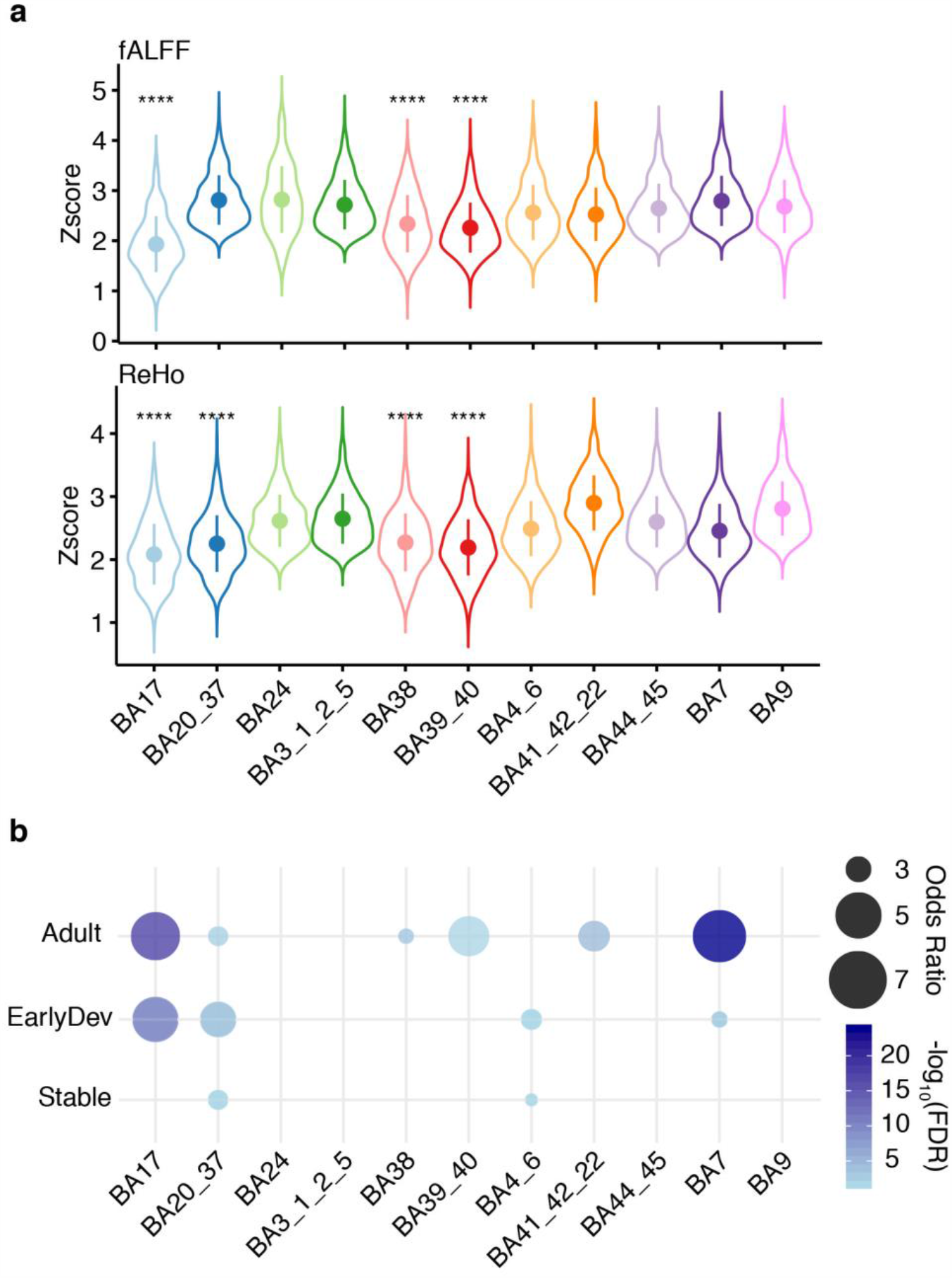
Leave one region out (LoRo) analysis underscores the importance of specific brain regions. **a**, Violin plots highlighting the contribution of each region in each rs-fMRI measurement. Violins represent the Z-score (Y-axis) of differential correlation after the specific region (X-axis) was removed. **** corresponds to a significant lower Z-score compared with other regions (p < 0.001; One-sided Wilcoxon’s rank sum test). Dots represent the mean Z-score for the specific Brodmann area. Lines represent the standard deviation (SD). N = 355. **b**, Bubblechart with -log_10_(FDR) and Odds Ratio from Fisher’s Exact test representing enrichment between developmental groups and genes differentially expressed in each region. X-axis shows abbreviations for each region. Y-axis represents each developmental cluster identified.

To further understand the functional properties of the rs-fMRI genes, we performed enrichment for gene expression data derived from single-cell RNA-seq studies^23^ (Methods). Intriguingly, we observed that the genes in the Adult cluster were highly enriched for parvalbumin (*PVALB*) expressing interneurons whereas EarlyDev genes were enriched for excitatory neurons. No cell-type enrichment was detected for the genes in the Stable cluster (Extended Data Fig. 6c). Because PVALB expression follows an anterior to posterior regional gradient^24^, we imputed *PVALB*^+^ interneurons abundance for each of the region analyzed (see Methods). As expected, the *PVALB*^+^ interneurons fractional abundance was higher in posterior regions compared with anterior regions (Extended Data Fig. 6d). Intriguingly, the relative abundance of these interneurons was significantly reduced in ASD posterior regions such as BA7 and BA17. These data indicate that our results are potentially driven by *PVALB*+ interneurons regional abundance further demonstrating the important role of these interneurons in ASD pathology. We next investigated the association of developmental gene clusters with genomic data from brain disorders including ASD^25^. The Adult cluster is enriched for downregulated genes in ASD and schizophrenia in line with our initial finding (Fig. 3a) while the EarlyDev cluster is enriched for upregulated genes in ASD (Extended Data Fig. 6e). This result was further confirmed using modules of co-expressed genes dysregulated in such disorders (Extended Data Fig. 6f). We next evaluated protein-protein interactions (PPI) between DC genes using STRING^26^. We found that the Adult and EarlyDev clusters were significantly enriched for PPI (Score = 400; FDR = 0.009 and FDR = 0.02, respectively), further underscoring that DC genes tend to act in the same pathways and that such pathways might be involved in functional connectivity (Fig. 3c). Together, these data extend the emerging picture of molecular pathways disrupted in ASD corresponding to rs-fMRI measurements^3,4,27,28^.

### The relationship of rs-fMRI and gene expression is altered at the brain region level

Due to the limited number of samples per ROI, we were not able to assess the association between brain activity and gene expression at a regional level. We overcame this limitation with a leave-one-region out (LoRo) approach inferring the contribution of each region in our results (see Methods). Briefly, by leaving one of the 11 regions out at a time, we were able to test whether the differential correlation was affected by one region or several specific regions. We calculated the z-score from the z-to-r Fisher transformation from each analysis (Supplementary Table 3). Interestingly, we observed a significant contribution from the primary visual cortex (BA17) to the DC genes using both fALFF and ReHo values (Fig. 4a). We next examined the enrichment of regional differential expressed genes (DEG; FDR < 0.05, |log_2_(FC)| > 0.3; see Methods) between ASD-CTL in DC genes. We explored whether any of the developmental gene clusters were enriched for specific regional DEG. Interestingly, we found the highest enrichment of Adult and EarlyDev cluster genes in cortical areas associated with vision and proprioception (BA17 and BA7) (Fig. 4b). Furthermore, we inspected regional developmental trajectories of DC genes in CTL samples and surprisingly found that DC genes follow a different trajectory in BA17 compared to other regions for all gene clusters (Extended Data Fig. 6). This indicates that DC genes might be functioning differently in BA17 during development. Taken together, these results support the emergent role of the visual cortex in ASD pathophysiology^29,30^.

## Discussion

Assessing gene expression in the brain permits a relevant examination of how biological pathways might be altered in the tissue of interest and connected to genetic predispositions. Moreover, functional imaging provides an important window into phenotypes associated with mental illness. Combining these approaches can help begin to bridge the gap between genes and behavior. Indeed, previous work has demonstrated correspondence between human brain gene expression and functional connectivity as measured by fMRI^3,4,27^. However, the studies using brain gene expression only used neurotypical populations.

Local brain activity measures such as ReHo and fALFF can assess neuronal connectivity and activity. When restricted to a specific image acquisition site and age range (e.g. children or adolescents), previous studies using ReHo and fALFF have found significant differences between CTL and ASD subjects in cortical regions but in different brain regions and directions^31,32,33-35^. However, protocol variability across sites can induce inconsistent findings in functional connectivity^36^. A quantitative meta-analysis indicated that only connectivity between the dorsal posterior cingulate cortex and the right medial paracentral lobule consistently differs between ASD and CTL subjects across sites and ages^37^; however, these regions were not available for tissue sampling in this study. Structural imaging studies have also indicated the difficulty in finding differences between ASD and CTL subjects when no age restriction is imposed^38-40^. In contrast to these age and site-restricted reports, our study includes ages from 5-64 years and data from 37 sites whose differences are retrospectively normalized and such differences with previous studies likely underlies our finding of few significant differences in brain activity between cases and controls.

We speculated that the expression of genes and their association with brain activity may underscore their potential relevance for any functional brain activity that is disrupted in ASD. In line with this, our results suggest that genes typically associated with rs-fMRI have lost their association when ASD genomics are included. These genes are important for brain development, regional differences, and excitatory/inhibitory identity. As previously reported, GABAergic signaling is disrupted across mouse models of ASD^41^ and GABA interneurons have a key role for cortical circuitry and plasticity^42-44^. Interestingly, genes highly expressed in the Adult gene cluster that are significantly associated with brain activity are overrepresented in a subpopulation of inhibitory interneurons expressing parvalbumin. In contrast, genes highly expressed in early development are overrepresented in excitatory neurons. In line with the role of parvalbumin neurons in normal brain circuitry and oscillations^42,45,46^, this distinct association might underscore the excitation-to-inhibition ratio imbalance in autism. Moreover, the relative abundance explained by the Adult genes of *PVALB*+ interneurons is significantly decreased in ASD. Because spatial *PVALB*+ expression covaries with rs-fMRI across regions^24^, we hypothesized that the relative increased abundance of these interneurons in the visual cortices and conversely the reduction shown in ASD explains the differential correlation in the specific subset of Adult genes. Therefore, these results further underscore the important role of parvalbumin interneurons in autism. We hypothesize that genes severely dysregulated in autism such as *SCN1B* or *KCNAB3* might additionally contribute to the excitation-to-inhibition ratio affecting normal network function and circuitry. Moreover, previous studies have shown that inhibitory neurons control visual response precision with increased activity leading to a sharpening of feature selectivity in mouse primary visual cortex^30^.

Additionally, multiple lines of evidence have indicated that individuals with ASD show slower switching between images in binocular rivalry^29,47-49^. Here, we provide evidence that regional brain expression influences the association between rs-fMRI values and gene expression, with the visual cortex as the major contributor to the variance explaining the rs-fMRI – gene association. Therefore, these results contribute to a consistent emerging role of the visual cortex in ASD pathology.

Finally, any functional interpretation of the genes identified should be made with caution. Here, we assessed the relationship between gene expression and rs-fMRI across cortical areas based on correlation, which is not necessarily evidence of causation. Moreover, functional imaging analysis of the ROIs used does not show large differences between neurotypical and individuals with autism. Larger sample sizes and improved parcellation of imaging and expression datasets may in the future allow for a more detailed investigation of these genes at the regional level increasing both specificity and sensitivity. Additionally, candidate genes should be further analyzed *in vivo* using model systems to provide a basic understanding of their effects on brain activity. In conclusion, we have established that autism pathology severely affects the relationship between gene expression and functional brain activity. Our results uncovered genes that are important for excitation-to-inhibition ratio balance and visual cortex function. These results provide molecular mechanisms for future studies relevant to understanding brain activity in individuals with autism.

## Data Availability

Custom R code and data to support the data correction, correlation analysis, visualizations, functional, and gene set enrichments are available at https://github.com/konopkalab/AUTISM_rsFMRI_GeneExpressionCorrelations

## Methods

### fALFF and ReHo

To provide image-derived phenotypes (IDPs) for each subject in the ABIDE cohort, regional measures of brain function were computed including the fractional amplitude of low-frequency fluctuation (fALFF) and regional homogeneity (ReHo). Extended Data Fig. 1 outlines and illustrates the main processing steps of the image analysis pipeline.

### Imaging Materials

This study used resting-state fMRI from the ASD and CTL subjects of both ABIDE I and ABIDE II^13,50^. After the removal of subjects with image artifacts or poor MNI152 coregistration, we analyzed the data from the remaining 916 ASD (86% male), and 1067 CTL (76% male) subjects, whose age ranges from 5-64 years.

### fMRI preprocessing

All data from each subject were preprocessed consistently—as described below—and are illustrated in Extended Data Fig. 1. We applied BETs 3dSkullStrip^51^ to remove both skull and non-brain tissue. To correct for possible head movement, volume realignment was applied by frame to frame, registering each volume to the volume preceding it with linear affine transformations using C-PAC (v1.7.1). We removed common brain fluctuations affecting the whole brain using global signal regression (GSR).

### Calculation of fALFF and ReHo

We computed fALFF and ReHo from the resting-state fMRI using C-PAC (v1.7.1)^52^ in native subject space, resulting in a volumetric map of fALFF and a map of ReHo for each subject. fALFF^15^ quantifies the slow oscillations in brain activity that form a fundamental feature of the resting brain. ALFF is defined as the total power within the low-frequency range (0.01 to 0.1 Hz) and forms an index of the intensity of the low-frequency oscillations. The normalized ALFF known as fALFF is defined as the power within that low-frequency range normalized by the total power in the entire detectable frequency range. fALFF characterizes the contribution of specific low-frequency oscillations to the entire frequency range^15^. To increase the signal to noise ratio by removing high-frequency information, we spatially smoothed each derivative map with a Gaussian kernel (FWHM=6mm).

ReHo^16^ aims to detect complementary brain activity manifest by clusters of voxels rather than single voxels as in fALFF. ReHo evaluates the similarity of the activity time courses of a given voxel to those of neighboring voxels using Kendall’s coefficient of concordance (KCC)^53^ as the index of time series similarity. This measure requires the cluster size as an input to define the size of the neighborhood. In this study, we used a cluster size of 27 voxels. The 26 neighbors of a voxel, x, are those within a 3×3×3 voxel cube centered on voxel x. The similarity of the activation time courses between each voxel, x, and it’s 26 nearest neighbors using 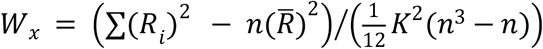 *W*_*x*_ is the KCC for voxel and ranges from 0 to 1, representing no concordance to complete concordance. *R*_*i*_ is the rank sum of the i-th time point. *R* is the mean value over the *R*_*i*_’s. K is the cluster size for the voxel time series (here K=27). n is the total number of ranks.

### Registration

The mean processed fMRI image was nonlinearly registered directly to an EPI template in MNI152 space using the symmetric normalization (SyN) non-linear registration method of the ANTs (v2.3.5) package^54,55^. The resulting transform was then applied to both the fALFF and ReHo maps to provide derivative maps in normalized MNI152 space. We used EPInorm-based registration as it better accounts for nonlinear B0 field inhomogeneities at the air to tissue interfaces^56,57^. Extended Data Figure 7 illustrates the improvement EPInorm-based registration has over more the commonly applied T1norm based registration. In this study EPInorm registration yields more accurate spatial normalization of the brains to the standard atlas space in which regional values are computed. Regions of improved registration include the sinuses which present air/tissue interfaces that induce non-linear distortions which are properly handled through EPInorm co-registration. EPInorm registration also has substantially lower standard deviation around the brain periphery across the 1,983 subjects assessed.

### Segmentation

In this study, we adapted the standard Brodmann publicly available through MRICron (v1.0.9) to form the 11 multi-area regions from which tissue samples were drawn from matched donor brains. Table 1 illustrates how we combined Brodmann areas to generate 11 regions that correspond with the RNA sequence data. We used the resulting 11 region atlas to assign a region label (parcellate) to each voxel in the fALFF and ReHo maps to enable computation of the mean regional fALFF and ReHo values for all subjects.

### Site Correction

We collected the ABIDE data across 30 different sites. These sites used MRI devices from different manufacturers (Siemens, Philips, GE) and used different MRI pulse sequences and patient protocols, which can cause differences in the absolute value of the fMRI acquired and can affect fALFF and ReHo values. As the mean fALFF and ReHo varied between sites, we applied a correction to minimize site differences. To suppress site differences, we calculated the mean value for the derivative maps in each region for each site.

### Derivative map normalization

To provide better inter-subject comparisons, we normalized regional fALFF and ReHo values to the weighted mean, weighted by the number of voxels for each region, over all of the regional values for each subject. To reduce the impact of confounders, we regressed out age, site, and sex by fitting a linear model to each and taking the residuals.

### RNA-seq processing and analysis

Dup15q patients were removed from the initial data^14^. Technical replicates were collapsed by the maximum expression value and maximum RNA integrity value. A total of 302 Control and 360 ASD were used for the final analysis. Only protein-coding genes were considered. Counts were normalized using counts per million reads (CPM) with the *edgeR* (v3.32.0) package in R^58^. Normalized data were log2 scaled with an offset of 1. Genes were considered expressed with log_2_(CPM+1) > 0.5 in at least 80% of the subjects. Normalized data were assessed for effects from known biological covariates (*Sex, Age, Ancestry, PMI*), technical variables related to sample processing (*Batch, BrainBank, RNA Integrity value (RIN)*) and technical variables related to sequencing processing based on PICARD statistics (https://broadinstitute.github.io/picard/).

We used the following sequencing covariates:

picard_gcbias.AT_DROPOUT, star.deletion_length, picard_rnaseq.PCT_INTERGENIC_BASES, picard_insert.MEDIAN_INSERT_SIZE, picard_alignment.PCT_CHIMERAS, picard_alignment.PCT_PF_READS_ALIGNED, star.multimapped_percent, picard_rnaseq.MEDIAN_5PRIME_BIAS, star.unmapped_other_percent, picard_rnaseq.PCT_USABLE_BASES, star.uniquely_mapped_percent.

Residualization was applied using a linear model. All covariates except Diagnosis, Subjects and Regions were taken into account:

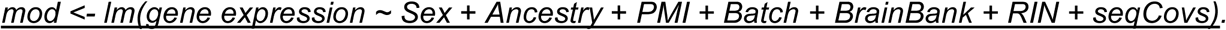

This method allowed us to remove variation explained by biological and technical covariates.

Adjusted expression was calculated extracting the residuals per each gene and adding the mean of the gene expression: *<u>adjusted</u> <u>gene</u> <u>expression</u> <u><-residuals(mod)</u> <u>+</u> <u>mean(gene</u> <u>expression)</u>*

Adjusted CPM values were used for rs-fMRI – gene expression correlation and resultant visualization.

### fMRI-gene expression correlation analysis

We performed Spearman’s rank correlation between the mean regional values of fALFF and ReHo and the regional gene expression across the 11 cortical areas analyzed. To define fMRI-gene expression relationships, we used random subsampling (200 times) of neurotypical individuals from the ABIDE I and II datasets. We matched the number of subjects per each cortical area (e.g. 25 ASD subjects for BA17). We performed correlation across the regions using all 11 areas matching with the gene expression dataset and averaged Spearman’s rank statistics over the 200 subsamples. P-values from Spearman’s rank statistics were adjusted by Benjamini-Hochberg FDR. Differential Correlation analysis was performed comparing the resulting Rho from neurotypical individuals to individuals with ASD for each gene using the *psych* (v2.0.12) package in R. Significant results are reported at FDR < 0.05 for neurotypical individuals statistics and P-value of differential correlation at p < 0.05. Differentially Correlated genes (DC genes) are based on the overlap between significant results in fALFF and ReHo.

### Leave-one-region out (LoRo) analysis

We performed the same subsampling approach followed by differential correlation analysis as described above leaving one region out at the time. This method allowed us to determine the effect of each region in the resultant z from the differential correlation analysis between healthy individuals and autistic individuals. Next, we calculated the contribution of each region based on a principal component analysis using the resultant z-values. We visualized resultant contributions in a multi-dimensional plot.

## Developmental analysis

The identification of gene clusters with different developmental trajectories was performed on DC genes using all subjects except for individuals above 60yr as they were represented only in the ASD group. We applied residualization as previously described removing the age from the covariates.

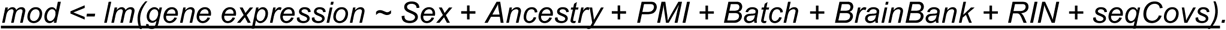

Then, we scaled gene expression and divided genes into 3 clusters according to the scaled expression values of healthy subjects only, using the *Kmeans* function from the *amap* (v0.8) package in R. We plotted the developmental trajectories using the loess regression and *ggplot2* (v3.3.2) package in R. To make loess regression computationally possible, 8000 data points were randomly sampled. Repeated samplings yielded very similar patterns. We made no adjustments for developmental time points and the x-axis directly represents the age of the subjects. We annotated clusters based on visual inspection of their trajectory. To subsample diagnosis-region groups (e.g ASD BA17 samples), we determined the diagnosis-region group with minimum number of samples and randomly subset other groups to that number. Then we plotted expression values with loess regression as before. To assess the significance of regional trajectories, we compared 0-10 years old and 10-20 years old samples in CTL. To equalize the sample number, we took the mean expression value per gene across all samples per age group. Then we applied t-test with FDR < 0.001 cutoff to decide significance. T-test was one-tailed for Adult (Ha: 0-10 < 10-20) and EarlyDev (Ha: 0-10 > 10-20) clusters but two-tailed for Stable cluster (Ha: 0-10 ≠ 10-20).

### Allen single nuclei RNA-seq analysis

Multi-Region snRNA-seq^23^ (MTG, V1C, M1C, CgGr, S1C, A1C) was from the Allen Brain Map portal (https://portal.brain-map.org/atlases-and-data/rnaseq). To identify cell-markers, we performed a differential expression analysis using Seurat^59^ (v3.9.9) with *FindMarkers* function based on Wilcoxon-rank sum test statistics. Markers were defined by Percentage of Cells expressing the gene in the cluster > 0.5, FDR < 0.05 and |log_2_(FC)| > 0.3.

### Functional Enrichment

We performed the functional annotation of differentially expressed and co-expressed genes using ToppGene^60^. We used the GO and KEGG databases. Pathways containing between 5 and 2000 genes were retained. We applied a Benjamini-Hochberg FDR (*P* < 0.05) as a multiple comparisons adjustment.

### Gene set enrichment

We performed gene set enrichment for neuropsychiatric DGE^25^, neuropsychiatric modules^25^, and cell-type markers^23^ using a Fisher’s exact test in R with the following parameters: alternative = “greater”, conf.level = 0.95. We reported Odds Ratios (OR) and Benjamini-Hochberg adjusted P-value (FDR).

### Protein-protein interaction analysis

We downloaded the STRINGdb (v2.2.0)^26^ in R and used it for the protein-protein interaction enrichment and annotation. Interactions with a score lower than 400 were removed. We performed visualization using *igraph* (v1.2.6) in R.

### Deconvolution

Deconvolution was performed by *MuSiC* (v0.1.1)^61^ in R. Single-cell data reference was downloaded from the Allen Brain Map portal (https://portal.brain-map.org/atlases-and-data/rnaseq).

### Code availability

Custom R code and data to support the data correction, correlation analysis, visualizations, functional, and gene set enrichments are available at https://github.com/konopkalab/AUTISM_rsFMRI.

### Statistical Analysis and Reproducibility

No statistical methods were used to pre-determine sample sizes. Nevertheless the data here reported is in line with the sample size of previous studies^62,63^. Samples were not randomized. ASD subjects with Chromosome 15q Duplication were excluded from the downstream analysis. Data collection and analysis were not performed blind to the conditions of the experiments. Findings were not replicated due to the limitation of the multi-region ASD transcriptome data. Nevertheless, we used two independent rs-fMRI measurement to refine and increase the confidence of our findings. For fALFF/ReHo rs-fMRI values and bulk RNA-seq transcriptomic data, distribution was assumed to be normal but this was not formally tested. Non-parametric tests have been used to avoid uncertainty when possible. Data collection and analysis were not performed blind to the conditions of the experiments.

### Reporting Summary

Further information on research design is available in the Life Sciences Reporting Summary linked to this article.

## Acknowledgments

G.K. is a Jon Heighten Scholar in Autism Research at UT Southwestern Medical Center. This work was supported by the NIMH (MH102603), NINDS (NS106447), and the James S. McDonnell Foundation 21^st^ Century Science Initiative in Understanding Human Cognition – Scholar Award (220020467) to G.K.

## Author Contributions

S.B., E.C., A.T., D.L., A.M., and G.K. analyzed the data and wrote the paper. J.H., M.J.G., and D.H.G. collected samples, processed RNA, and generated bulk RNA-seq libraries. A.T. analyzed the ABIDE I and ABIDE II data. A.M. and G.K. designed and supervised the study, and provided intellectual guidance. All authors discussed the results and commented on the manuscript.

---

**Extended Data Figure 1:**
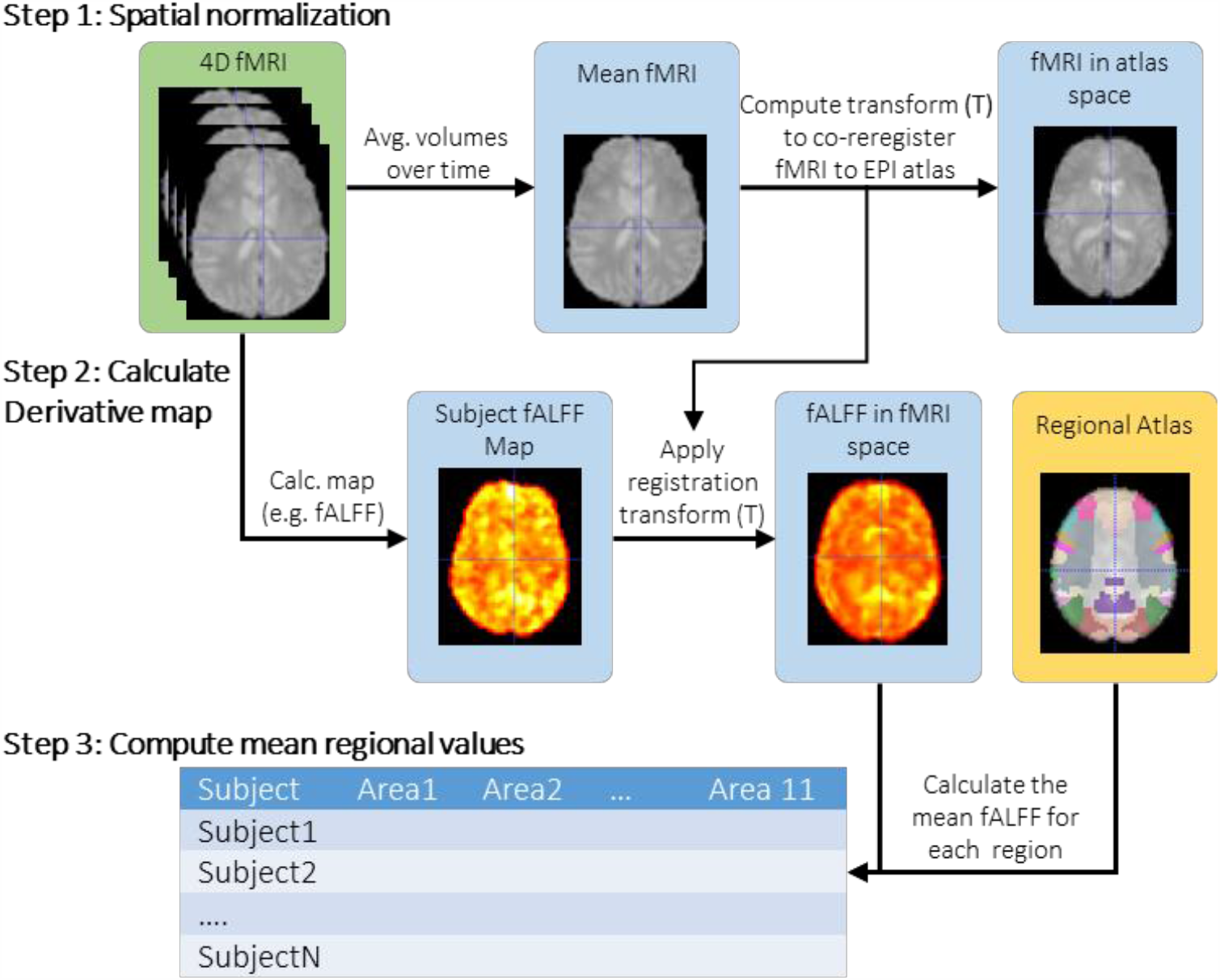
Image analysis pipeline. Three steps are used to calculate regional values from MRI. Step 1: Subject fMRI (green) is spatially normalized to atlas space. Step 2: Local functional activity measures are derived for each subject (e.g. fALFF) and coregistered to atlas space. Step 3: Mean regional measures are computed using the atlas (yellow) for each region for each subject.

**Extended Data Figure 2.**
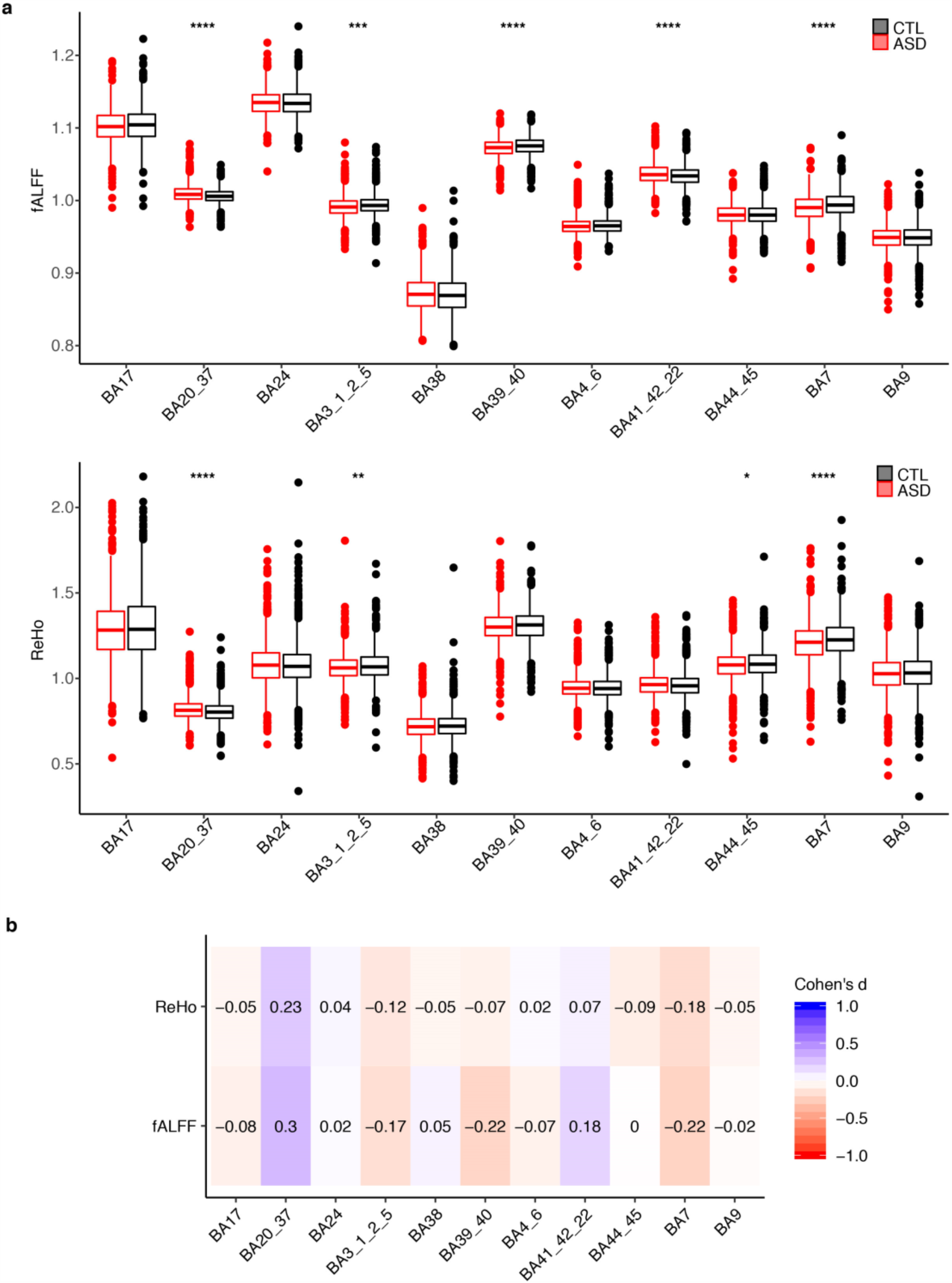
rs-fMRI data quality control. **a**, Boxplots of fALFF and ReHo measurements comparison between ASD (red) and CTL (black) across multiple ROIs analyzed. Stars correspond to the significant differences between ASD and CTL based on one-sided Wilcoxon rank sum’s test (**** p < 0.001, *** p = 0.01, * p = 0.05). Boxes extend from the 25th to the 75th percentiles, the center lines represent the median. **b**, Heatmap of Cohen’s D (effect sizes) derived from ASD – CTL comparison for both rs-fMRI measurements across the ROIs analyzed.

**Extended Data Figure 3.**
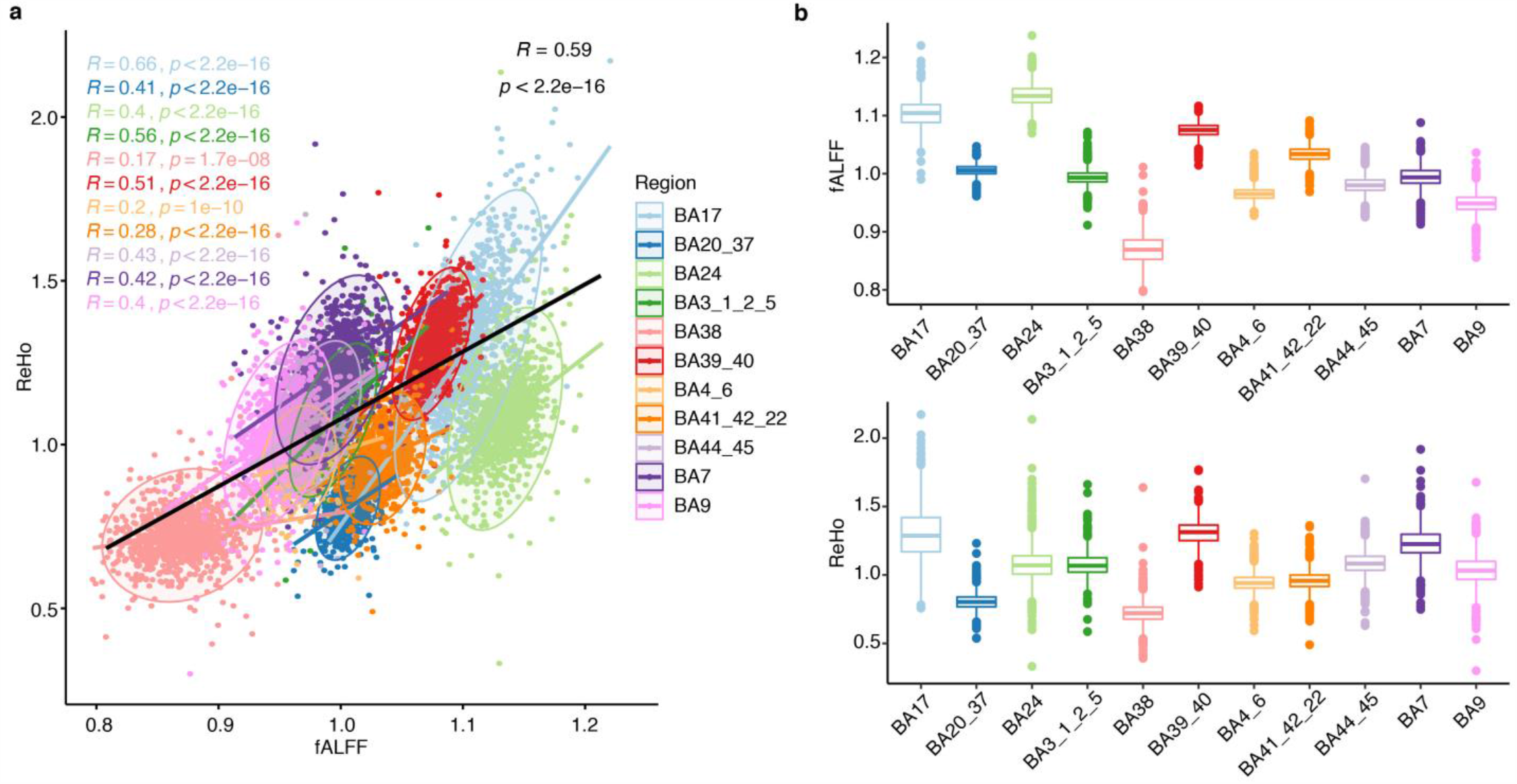
**a**, Scatter plot comparing fALFF (X-axis) and ReHo (Y-axis) between the 11 ROIs analyzed in CTL. Spearman rank *rho* values and associated p-values are shown colored by ROIs. Black line corresponds to the across-ROIs (pancortical) correlation. **b**, Distribution of fALFF and ReHo measurements in the 11 ROIs analyzed in CTL. Boxes extend from the 25th to the 75th percentiles, the center lines represent the median.

**Extended Data Figure 4.**
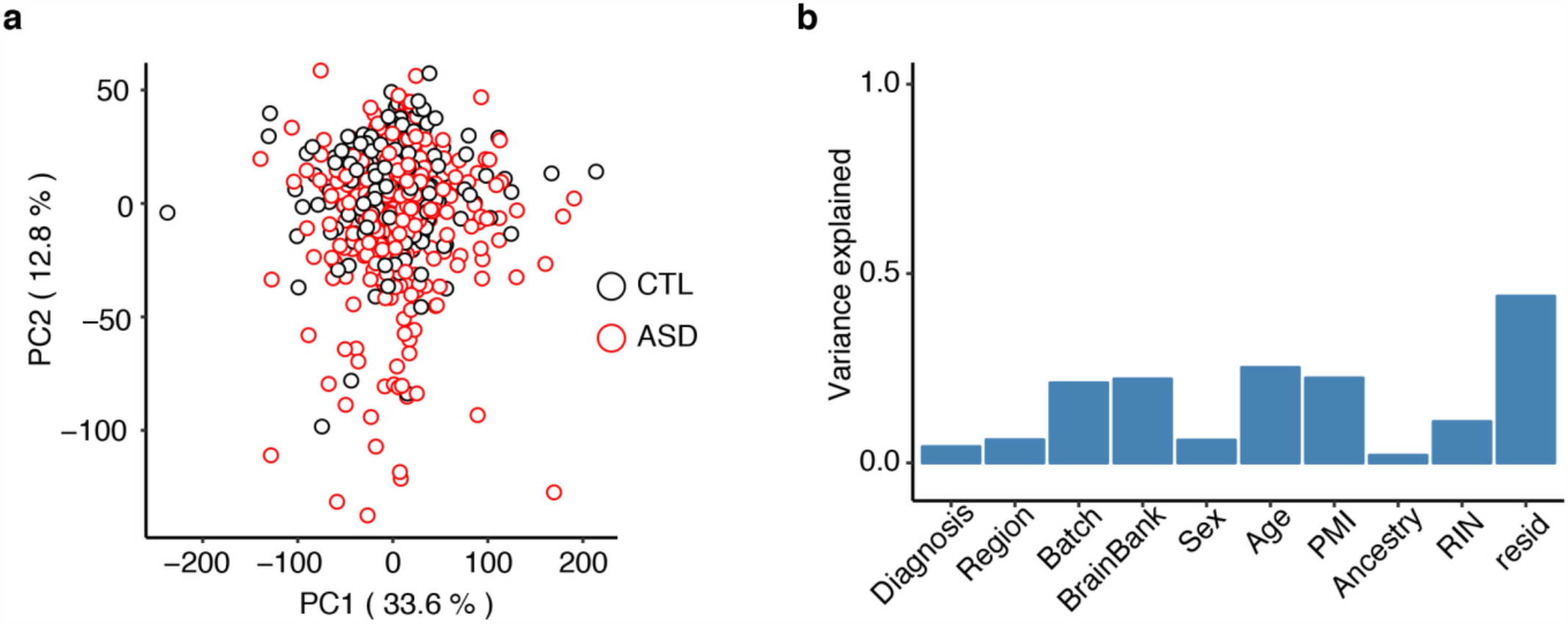
**a**, Principal component analysis based on the RNA-seq data of all the subjects used in this study. **b**, Variance explained by each covariate adjusted across 10 principal components.

**Extended Data Figure 5.**
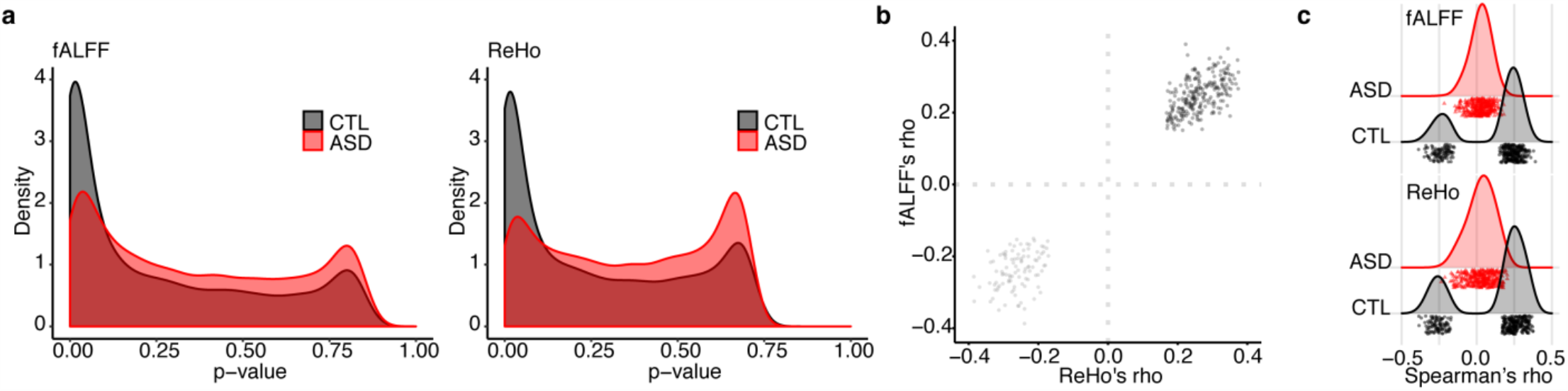
**a**, Distribution of Spearman’s rank p-values in both rs-fMRI measurements for ASD and CTL. **b-c**, Scatterplot of the comparison between CTL *rho* values for the differentially correlated genes (DC genes) and (**c**) the comparison between ASD and CTL for both fALFF and ReHo.

**Extended Data Figure 6.**
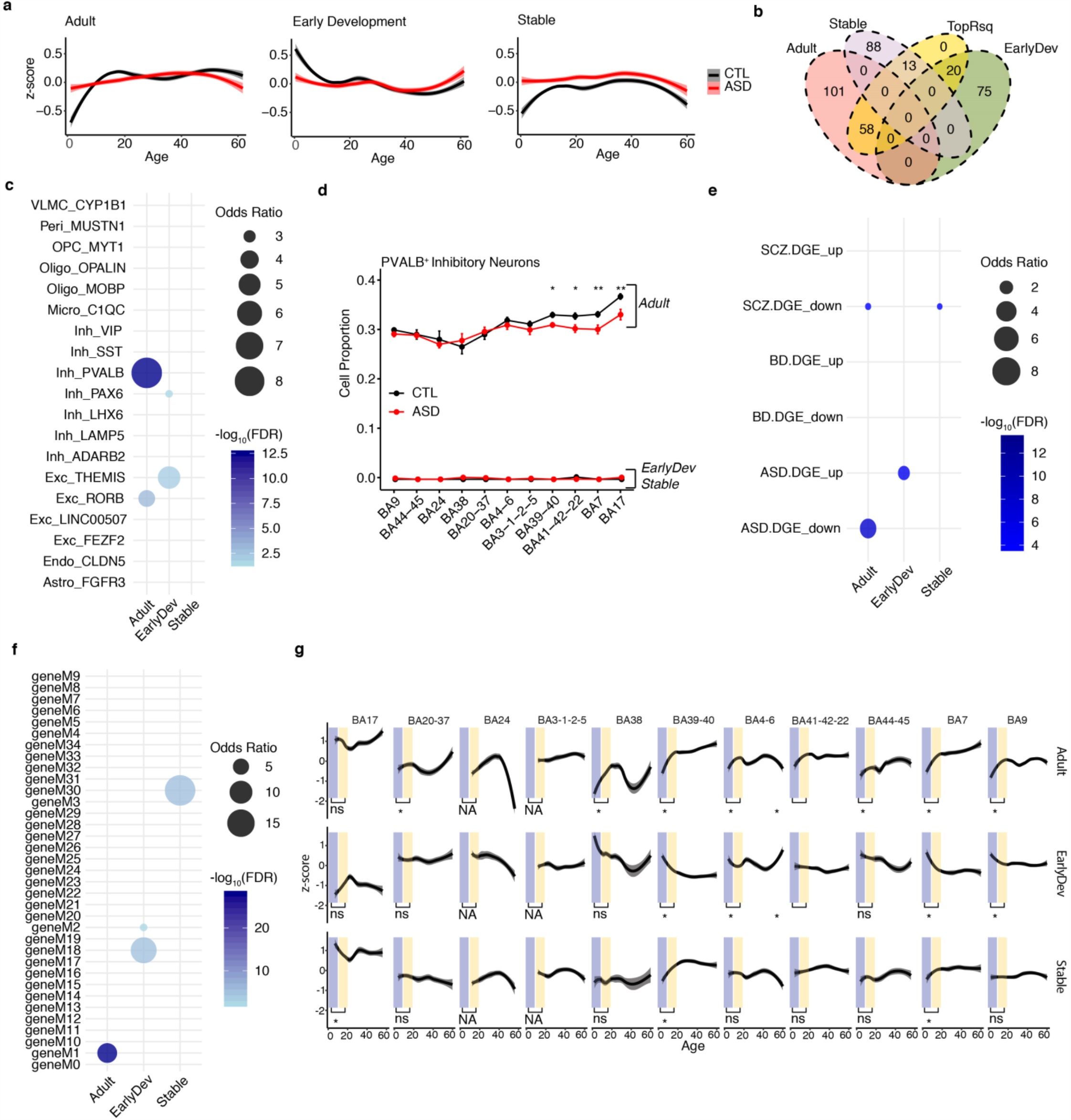
**a**, Developmental trajectories after equalizing diagnosis-region groups by random subsampling. Loess regression was used to fit smooth curve for the values of all genes per cluster across development. Smooth curves are shown with 95% confidence bands. Y-axis indicates Z-scored gene expression. **b**, Overlap between the developmental clusters and the TopRsq genes. **c**, Bubble-chart of enrichment between developmental clusters (X-axis) and cell-type markers (Y-axis) calculated using single-nuclear RNA-seq from temporal cortex. **d**, Line chart showing the median with standard error of PVALB+ interneurons imputed proportions across the 11 regions analyzed based on Adult, EarlyDev, Stable genes. Stars correspond to the significant differences between ASD and CTL based on one-sided Wilcoxon rank sum’s test (** p = 0.01, * p = 0.05). Y-axis indicates cortical regions by anterior-to-posterior ordering. **e**, Bubble-chart of enrichment between developmental clusters (X-axis) and genes differentially regulated in autism, bipolar disorder, and schizophrenia (Y-axis) from an independent study. **f**, Bubble-chart of enrichment between developmental clusters (X-axis) and modules associated with autism, bipolar disorder, and schizophrenia (Y-axis) from an independent study. **g**, Statistical analysis on trajectories of gene clusters in CTL samples across regions. Blue shades indicate 0-10 years old, yellow shades indicate 10-20 years old and asterisks indicate significance (FDR < 0.001 in t-test; one-tailed for Adult and EarlyDev, two-tailed for Stable cluster). Y-axis indicates Z-scored gene expression. X-axis indicates the age. Loess regression with confidence intervals is over imposed to depict the overall distribution. Smooth curves are shown with 95% confidence bands.

**Extended Data Figure 7:**
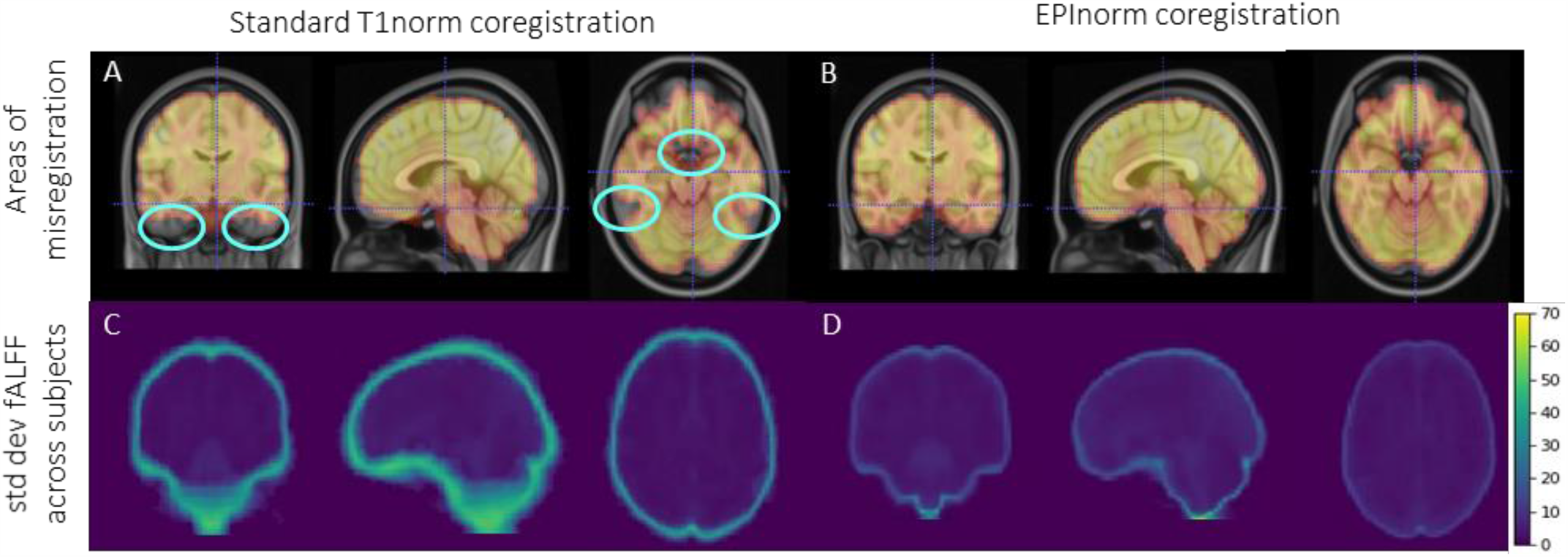
Quantitative comparison of EPInorm-based versus T1norm-based co-registration. Each panel shows a mid-coronal image (left), mid-sagittal image (middle) and mid-axial image (right). Top panels show fMRI (yellow/red overlay) coregistered to MNI T1 anatomical atlas (underlay) using (A) T1norm-based co-registration and (B) EPI based registration. Both co-registrations are 3-dimensional. EPInorm based co-registration better aligns the derivative maps (fALFF and ReHo) to brain anatomy and reduces areas of misregistration (blue circles). Bottom panels show the standard deviation for fALFF values across subjects using (C) T1norm-based co-registration and (D) EPInorm-based co-registration.

